# Evaluation of the Panbio^™^ rapid antigen test for COVID-19 diagnosis in symptomatic health care workers

**DOI:** 10.1101/2021.06.21.21259234

**Authors:** Marjan J. Bruins, Claudy Oliveira dos Santos, Marjan Spoelman-Lunsche, Marieke I. van den Bos-Kromhout, Sylvia B. Debast

## Abstract

If a health care workers (HCW) experiences COVID-19 associated symptoms, SARS-CoV-2 testing must be performed as soon as possible, to prevent transmission of the virus and to guarantee continuity of care. The gold standard for the detection of SARS-CoV-2, RT-PCR, has a high sensitivity but usually takes 6-8 hours. Lateral flow antigen assays take only 15-30 minutes and do not need any high tech equipment.

In a prospective study of our hospital’s HCWs, we evaluated the sensitivity of the Panbio™ COVID-19 Ag Rapid Test (Abbott) against the molecular test Aptima™ SARS-CoV-2 Assay (Hologic) which uses Transcription Mediated Amplification (TMA). TMA positive samples were further subjected to a quantitative real-time SARS-CoV-2 PCR to obtain Ct values as an indication of the viral load.

Of 1101 HCWs included in the study between November 2020 and February 2021, 84 (7.6%) were TMA positive, of which 48 (57.1%) were antigen test positive. Most false negative antigen test results occurred if the duration of symptoms had been ≤1 day or ≥7 days. Sensitivities for symptom onset of ≤1, 2 or 3 days were 47.1%, 63.0% and 66.7% respectively.

The Panbio™ rapid test is fast and easy to perform, but is not a suitable SARS-CoV-2 test to confirm or exclude COVID-19 in HCWs with a very recent onset of symptoms.

## Introduction

The COVID-19 pandemic that has ruled the world since March 2020, is a global burden on all health care systems. Hospitals and their emergency services and intensive care units have been straining to cope with the high number of admissions and the severity of the disease with its subsequent high mortality rate^1^.

Health care workers (HCWs), especially those in direct contact with patients, are at an increased risk of SARS-CoV-2 infection^2^. To prevent hospital transmission and to maintain the workforce, rapid testing of HCWs with COVID-19 associated symptoms is extremely important.

Nucleic Acid Amplification Tests (NAAT, e.g. PCR) are the gold standard for the detection of SARS-CoV-2. Most assays have a high sensitivity, but need advanced instrumentation and training, and take several hours to complete. Recently developed lateral flow antigen assays for the detection of COVID-19 take only 15-30 minutes, do not need any additional high tech equipment, are suited for point-of-care testing and cheaper than PCR-based rapid tests. Because rapid antigen tests are less sensitive than molecular assays, they are expected to perform best when viral loads are high, until approximately seven days after onset of symptoms^3,4,5^. To obtain a reliable result, a rapid test with a sensitivity of at least 80% and a specificity higher than 97% is recommended^6^.

If a rapid test can exclude COVID-19 in HCWs with COVID like symptoms, their absence while awaiting test results is reduced to a minimum amount of time and continuity of care can be guaranteed. However, because HCWs are preferably tested immediately as soon as symptoms appear, the question is whether an antigen test is sensitive enough to yield a positive test result^3^.

In a prospective study, we evaluated the use of the Abbott Panbio™ COVID-19 Ag Rapid Test, to exclude or confirm COVID-19 in our hospital’s HCWs and to determine its sensitivity for this specific group with (mostly) early symptoms.

## Study design

### Setting

According to our hospital’s policy, HCWs experiencing one or more COVID-19 associated symptoms (cough, runny or stuffy nose, sore throat, loss of sense of smell or taste, shortness of breath, myalgia, fever) were expected to have themselves tested as soon as possible. Through an online appointment system, they could be tested the same day at our hospital sampling site. A combined nasopharyngeal/throat swab was taken by trained personel with a FLOQSwab® (Copan Diagnostics, Inc., Brescia, Italy) and transported in Hologic lysis buffer (Hologic, San Diego, CA, USA) to the clinical microbiology laboratory, where a SARS-CoV-2 NAAT was performed. Within six to twelve hours of sampling, the test result was reported to the HCW. If the NAAT was positive, an infection control practitioner instructed the HCW about the prevailing quarantine rules. If negative, the HCW could return to work, provided the illness permitted this.

### Sample size

An earlier evaluation of the Panbio™ Rapid Test among symptomatic subjects reported a sensitivity of 72.6% and a specificity of 100%^7^. Based on a prevalence of 8% measured among our hospital’s HCWs during the first COVID-19 peak in March 2020 and an expected sensitivity of 70%, approximately 1,000 samples were needed to determine a specificity of 99% with a 95% confidence interval (CI) of 10% width^8^.

### Ethics approval and informed consent

The local Medical Research Ethics Committee considered the study not subject to the Dutch Medical Research Involving Human Subjects Act (WMO) and declared it to be exempted from further review. All participants signed a written informed consent form.

### Panbio™ COVID-19 Ag Rapid Test

The Panbio™ COVID-19 Ag Rapid Test (Abbott, Lake Country, IL, USA) is a lateral flow assay which detects SARS-CoV-2 nucleocapsid protein in nasopharyngeal specimens. The sample taken with the swab provided in the Panbio™ test kit is inserted in a tube with 300 µl extraction buffer, swirled in the fluid and squeezed out. Five drops of suspension are dispensed into the sample well of the test cassette. The sample migrates along the membrane in the cassette, where virus antigen-specific conjugate complex binds to antibodies on the test line, which turns red. A non-specific conjugate binds to matching antibodies in the control section, producing a red line indicating test validity. The test is read after 15-20 minutes. Any visible test line means a positive test result.

### SARS-CoV-2 NAATs

The qualitative SARS-CoV-2 NAAT used was the Aptima™ SARS-CoV-2 Assay performed with the Panther Fusion® System (Hologic, San Diego, CA, USA). This NAAT combines the technologies of target capture, Transcription Mediated Amplification (TMA), and Dual Kinetic Assay (DKA).

In TMA positive samples the viral load was determined with an in-house developed quantitative real-time SARS-CoV-2 PCR, using a KingFisher Flex extractor (Thermo Fisher Scientific, Waltham, MA, USA) on an Applied Biosystems® 7500 Real-Time PCR System platform (Thermo Fisher Scientific)^9^.

### Participants and data analysis

From all HCWs who came for SARS-CoV-2 testing and who gave informed consent, a second nasopharyngeal specimen was taken. A trained laboratory technician performed the Panbio test on site immediately after sampling according to the manufacturer’s instructions. Test results were read and recorded after 15 minutes of incubation at room temperature and test cassettes were photographed for later reference. For each participant we registered the duration of symptoms. Sensitivity and specificity with 95% confidence intervals (Cis) of the antigen test were calculated in comparison with the qualitative Aptima™ results. Quantitative PCR was performed on positive samples to obtain cycle threshold (Ct) values.

## Results

In total 1,101 SARS-CoV-2 tests were performed, from 1,014 HCWs of whom 869 (86%) were women. The median age was 39 years (range 17-66 years). Of 1,101 tests, 84 were PCR positive (prevalence of 7.6%). Of these, 48 were positive in the Panbio™ COVID-19 Ag Rapid Test, yielding an overall sensitivity of 57.1% (95% CI 45.9-67.9%). Table 1 shows the results per symptom duration. If the symptoms had started within the last 24 hours, sensitivity dropped to 47.1% (95% CI 23.0-72.2%). The highest sensitivity was reached after three and four days after symptom onset. Of 77 NAAT positive samples the Ct value could be determined, which ranged from 8.8 to 37.6. Three samples could not be tested and four Aptima™ positive samples were negative in the quantitative PCR.

**Table 1.** Test results of NAAT and Panbio test per duration of symptom onset.

| Symptom onset (days) | N | NAAT positive n | Panbio test positive n | Average Ct value NAAT | Average Ct value Panbio | Sensitivity Panbio (%) |
| --- | --- | --- | --- | --- | --- | --- |
| ≤ 1 | 295 | 17 | 8 | 24.8 | 20.9 | 47.1 |
| 2 | 593 | 46 | 29 | 21.5 | 20.5 | 63.0 |
| 3 | 128 | 12 | 8 | 24.1 | 19.2 | 66.7 |
| 4 | 85 | 3 | 2 | 23.0 | 19.9 | 66.7 |
| 5 | 15 | 2 | 1 | 22.9 | 16.6 | 50.0 |
| 6 | 14 | 0 | 0 | - | - | - |
| ≥ 7 | 33 | 4 | 0 | 34.0 | - | 0.0 |
| Total | 1101 | 84 | 48 |  |  | 57.1 |

Figure 1 shows the results of the antigen test in relation to PCR Ct values and duration of symptoms. Most false negative antigen test results occurred in HCWs with low viral load (Ct >30), consistent with early (less than 24 hours) or longer lasting (seven days or more) symptoms. The antigen test was easy to perform and read. There were no inconclusive or false positive results. Specificity was 100%.

**Figure 1.**
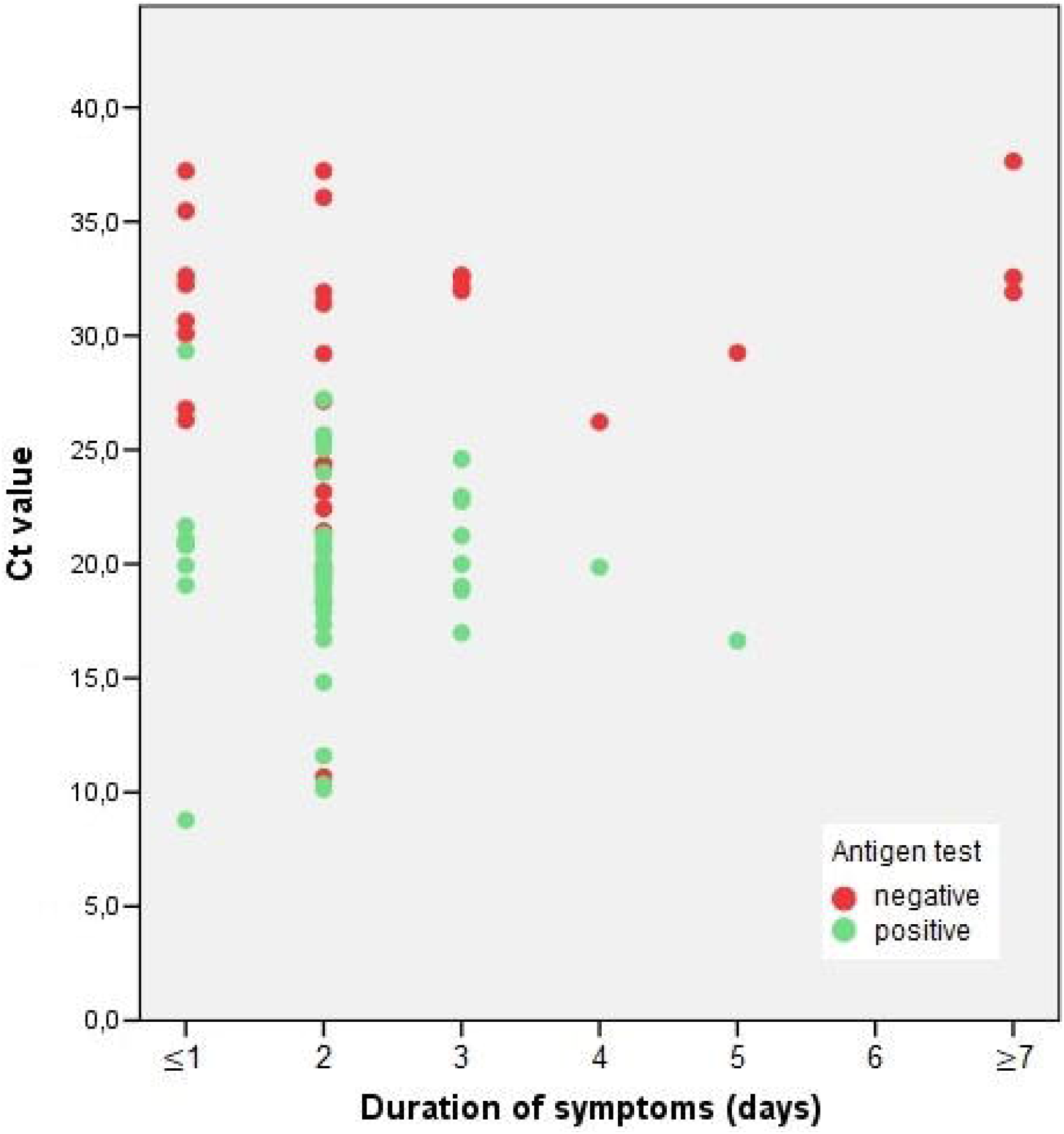
Results of the Panbio™ antigen test in relation to PCR Ct values and duration of symptoms.

## Discussion

In our study, the Panbio™ COVID-19 Ag Rapid Test was mainly false negative compared to NAAT during early and late stages of infection amongst HCWs. If COVID-19 symptoms had started within the last 24 hours, 27% (295/1101) of all participants tested false negative, sensitivity was less than 50%.

Results of the qualitative Aptima™ SARS-CoV-2 assay are based on DKA and light emission from labeled amplicons of specific regions of SARS-CoV-2 RNA, reported as Relative Light Units (RLU). The four samples that were Aptima™ positive and quantitative PCR negative had RLU values between 700 and 950, considered to be near the limit of detection (weakly positive samples). Partial degradation of virus RNA during storage can also have been the reason why no Ct value could be obtained.

One sample with Ct value 10.7 was negative in the Panbio™ test. Repeat antigen testing was negative, repeat CT value was 17.2. This discrepant result may have been due to the high mucous content of the sample.

In the beginning of COVID-19 infection, virus levels are still low, consistent with a high PCR Ct value. As our study shows, early after onset of symptoms, a SARS-CoV-2 antigen test may yield a false negative test result, which is inconclusive about the stage of the disease and does not preclude developing contagiousness. A limitation of our study was that we were not able to actively follow-up on symptomatic HCWs who tested Aptima positive and antigen test negative, to see whether the antigen test would become positive in the following days. Still, 81 HCWs were tested with both NAAT and antigen test more than once, of whom 21 made a new appointment because symptoms persisted or worsened For these however, test results of first and repeat sampling were all negative, except for one HCW who tested Aptima positive and antigen test negative in the first round (and negative for both in the second tests), probably due to a much earlier acquired, previous infection (symptoms onset 28 days ago, Ct value 32.6).

In other studies evaluating the Panbio™ COVID-19 Ag Rapid test, sensitivity ranging from 72% to 91% was found among symptomatic patients^7,10,11,12,13,14,15^. Specificity was almost always 100%. Sensitivities increased to over 90% at a Ct value of lower than 24, representing a peak viral load in the first week of infection and presumably indicating a limit of infectiousness^16^. This confirms the usefulness of the rapid antigen test for diagnosing COVID-19 in a high prevalence population with symptom onset of several days, according to the WHO’s recommendations^6^. In previous studies however, the relation between duration of symptoms and antigen test results was not assessed for subjects with symptoms onset of ≤ 24 hours^10,15,17^. To our knowledge, ours is the first study evaluating this among HCWs. The short turnaround time and easy performance of the assay are major advantages, but the sensitivity proved to be too low for it to be used as a single point diagnostic test for timely identifying COVID-19 positive HCWs and prevent healthcare associated transmission.

## Data Availability

All data are available on request.

## Acknowledgements

We thank Marianne Egbers, Gerloes Komdeur, Marieke Kral, Saskia Schipper-van Dijk, Miranda Veltman-Wolbink, the sampling crew and all participants for their contribution to the study.

